# Projecting EMS Workforce Demand in an Aging State: A Florida Forecast Through 2035

**DOI:** 10.64898/2026.07.07.26357403

**Authors:** Bruce J. Moeller, Michael Lozano, Lindsay J. Peterson, Mohammad Al Olaimat, Mingyang Li, Alina Hagen, Hongdao Meng

## Abstract

**OBJECTIVES:** Population aging is a major contributor to increasing demand for emergency medical services (EMS), yet EMS workforce projections based on population data remain limited. This study projected future EMS incident volume and clinician workforce requirements in Florida from 2026 to 2035 based on historical data on EMS response records to inform workforce planning.

**METHODS:** We conducted a retrospective, population-based secondary analysis and forecasting study using de-identified state-wide emergency EMS response records from Florida’s Emergency Medical Services Tracking and Reporting System (EMSTARS) spanning January 1, 2017 through December 31, 2025. Incidents were assigned to seven age cohorts and aggregated into monthly time series. We used Seasonal Autoregressive Integrated Moving Average models with exogenous inputs (SARIMAX) to project age and cohort-specific incident volume for 2026 through 2035. Projected future incident volumes were translated into EMT and paramedic full-time equivalent (FTE) requirements using observed EMSTARS staffing configurations and target operational parameters.

**RESULTS:** Annual EMS incidents increased from 4.10 million in 2017 to 5.22 million in 2025 and are projected to reach 7.76 million by 2035, a 48.8% increase over the 2025 baseline. By 2035, adults aged 60 and older are projected to represent 31.2% of Florida’s population while accounting for 61.6% of all EMS incidents. Total estimated EMS workforce requirements are projected to increase from 9,542 FTEs in 2025 to 14,195 FTEs by 2035, requiring approximately 4,654 additional FTEs (a 48.8% increase).

**CONCLUSIONS:** Florida’s aging population is projected to drive a nearly 50% increase in EMS incident volume and associated workforce requirements over the next decade, with demand disproportionately concentrated among older adults. With a substantial concentration of adults aged 80 and older and a rapidly expanding oldest-old cohort, Florida is confronting the demographic conditions projected to emerge in other states over the next decade. The findings offer researchers and policymakers a replicable framework and a directly applicable planning reference for jurisdictions across the United States.

## Introduction

The U.S. population is aging rapidly, with the fastest growth in the oldest age cohorts; including adults aged 80 and older, referred to here as the oldest old; representing a major driver of demand for social and medical services, especially for Emergency Medical Services (EMS) (1–3). This major demographic shift has significant implications for health care delivery and workforce planning (4). Many older adults live with multiple chronic conditions, functional limitations, and cognitive impairment, increasing their likelihood of time-sensitive, unplanned care needs (5). These challenges are compounded by fragmented care delivery, limited geriatric expertise, and barriers to home and community-based services (4, 6, 7). For EMS, these forces raise not only clinical questions but also urgent operational questions about staffing capacity, unit availability, and long-term workforce requirements.

EMS systems serve as a critical safety net for older adults. Utilization increases substantially with age. Older adults use EMS at rates far exceeding those of younger populations and are more likely to require emergency department evaluation and hospitalization after EMS contact (8–10). This pattern is most pronounced among the oldest old, who account for a disproportionate share of prehospital encounters despite comprising a relatively small fraction of the overall population (8). As this cohort expands, age-specific utilization patterns will translate directly into higher incident volumes.

Advanced age is also associated with more complex prehospital assessment and disposition decision-making, which can increase cognitive workload and operational complexity for EMS clinicians and underscores the need for geriatric-informed clinical pathways and decision support (10, 11). Recurrent EMS use among older adults further underscores the ongoing burden on EMS systems and the potential for targeted strategies to address high-frequency users and care transitions (12). Together, increased volume and case complexity have implications for unit availability and the overall capacity of EMS systems to meet community needs. While we know population aging is accelerating demand for emergency medical services (EMS), EMS leaders have limited evidence to substantially guide and support forward-looking workforce planning. This study projects EMS incident volume and clinician workforce needs in Florida through 2035, integrating age-stratified utilization patterns, validated population projections, and operational staffing assumptions to anticipate EMS demand shifts that many other states are likely to face in the future.

## Methods

### Study Design and Regulatory Determination

We conducted a retrospective, population-based, secondary analysis and forecasting study to estimate EMS incident volume and associated EMT and paramedic workforce requirements in Florida through 2035. The analysis used de-identified EMS emergency response records from the Emergency Medical Services Tracking and Reporting System (EMSTARS), Florida population estimates and projections, and workforce capacity assumptions. The final analytic dataset comprised Florida 9-1-1 EMS responses from January 1, 2017, through December 31, 2025. This study period was selected to capture the longest available period of consistent statewide EMSTARS data and to improve estimation of temporal trends, seasonality, and age-specific utilization patterns.

The University of South Florida Institutional Review Board determined that the study meets the criteria for exemption from IRB review. EMSTARS data were provided under a data use agreement with the Florida Department of Health. No direct identifiers or protected health information were accessed. Workforce requirements were estimated using EMSTARS staffing configurations and prespecified full-time equivalent (FTE) capacity assumptions. All data were stored on secure University of South Florida-managed resources.

### Data Sources and Study Period

EMS response data were obtained from EMSTARS, Florida’s statewide EMS data system. The analytic dataset included de-identified record-level data for Florida 9-1-1 EMS responses from January 1, 2017, through December 31, 2025. From these records, incident date and time, patient age, dispatch timestamp, service completion or unit back-in-service time, and staffing configuration variables, including the number and type of EMS responders associated with each incident, were extracted. These incident-level variables were used to assign records to age cohorts, aggregate incidents into monthly time series for forecasting, and estimate observed staffing patterns for subsequent workforce calculations. To account for the gradual increase in onboarding rates in the EMSTARS over time, the incident-level digital records were supplemented with summary reports that agencies were required to submit to Florida’s Department of Health if they were not submitting electronic patient care reports (ePCR) to EMSTARS. These summary reports provided aggregate incident counts, which allowed for an assessment of the full EMS workload across all reporting agencies in the state, but other elements, such as age and dispatch times were not provided as part of these paper submittals. During the period from 2017 through 2025, these summary reports accounted for only 5.91% of all EMS events. These paper-based incident counts were included in the forecasting and scaling procedures described below.

Population estimates and projections were obtained from the University of Florida Bureau of Economic and Business Research (BEBR), which provides Florida’s official population estimates and projections by county, age, and demographic group. Population data were organized by age cohort and used to characterize demographic change, calculate age-specific EMS utilization patterns, and support projections of future EMS incident volume through 2035. Where population estimates or projections were unavailable for intermediate months or years, values were linearly interpolated to create a continuous time series aligned with the monthly EMS incident data.

### EMS Incident Variables

Incident-level EMSTARS records were used to derive variables for age-stratified forecasting and workforce estimation, with patient age used to assign incidents to age cohorts. For analysis and reporting, age groups were organized to distinguish younger populations from older adult cohorts, with particular attention to adults aged 60-69, 70-79, and 80 years and older. Records with documented patient age were included in age-cohort-specific modeling. Records with missing age, representing 18.19% of EMSTARS records, were not assigned to age cohorts but were accounted for in the missing-age adjustment described below.

For each EMS response, we extracted incident date and time variables to assign records to calendar month and year. Dispatch timestamp and service completion or unit back-in-service time were used to calculate incident duration in hours. Incident duration served as a workload input for FTE estimation. Records with implausible or invalid duration values were excluded from duration-based workforce calculations; specifically, values less than 0.1 hours or greater than 12 hours were excluded to reduce the influence of data entry errors and atypical cases.

Staffing configuration variables were extracted from EMSTARS records to determine the number and type of EMS responders for each incident. For this analysis, EMS clinicians were categorized as emergency medical technicians (EMTs) and paramedics (PMs). These fields were used to estimate the average number of EMTs and PMs per incident during the historical study period. The observed average staffing patterns were then applied to projected annual incident volume to estimate future workload hours separately for EMTs and PMs.

Incident records were aggregated by month and age cohort for forecasting. Monthly age-cohort incident counts served as the basis for the Seasonal Autoregressive Integrated Moving Average models with exogenous inputs (SARIMAX) described below. Annual incident totals were calculated by summing projected monthly counts for each calendar year. These annual projections were then used in the workforce FTE estimation framework.

### Population Data and Projection Inputs

For this analysis, BEBR population data were aggregated into the same age cohorts used in the EMS forecasting models. Population projections were aligned with the EMS age cohorts used in the SARIMAX analysis. The models forecasted EMS incident counts by age group; population data were used to characterize demographic change and support age-specific demand projections. Because BEBR population projections were unavailable for several intermediate years (e.g., 2024, 2026–2029, and 2031–2034), linear interpolation was performed to estimate the missing annual population values. These annual population estimates were aggregated into the EMS age cohorts and aligned with the monthly EMS incident series for demographic interpretation and age-specific demand projections.

Age-specific population denominators were used to calculate EMS incident rates per 1,000 population and to evaluate changes in utilization across age cohorts. Older adult cohorts were analyzed separately to assess whether projected EMS demand differed among adults aged 60-69 years, 70-79 years, and 80 years and older.

### Forecasting and Workforce Estimation Framework

We used a two-stage analytic framework to estimate future EMS incident volume and associated EMT and paramedic workforce requirements in Florida through 2035, with Stage 1 generating age-specific incident forecasts and Stage 2 translating these into EMT and paramedic FTE needs. Sensitivity analyses and model assumptions are described after the primary forecasting and workforce estimation procedures.

### Stage 1: SARIMAX Incident Forecasting

In Stage 1, age-specific EMS incident forecasts were generated using SARIMAX applied to monthly time series derived from incident-level EMSTARS data. Records were assigned to age cohorts and aggregated, preserving age-specific utilization patterns while capturing temporal trends and seasonality. A baseline model using incidents with documented patient age was estimated to produce age-specific forecasts and inform interpretation. Monthly projections were aggregated to annual totals for reporting and workforce estimation.

The adjusted model used a 2017–2025 analytic window and incorporated exogenous variables for reporting completeness and COVID-era disruption, including aggregate reporting percentage, reporting coverage ratio, and a binary indicator for 2020–2021. SARIMAX was selected given temporal autocorrelation and seasonality in EMS volume. Forecasts were generated through 2035, and monthly projections were aggregated to annual incident totals by age cohort.

### Reporting Completeness, Missing Age, and COVID-Era Adjustment

Because EMS reporting completeness and patient age documentation varied over the study period, we implemented procedures to reduce bias from missing age data and changes in reporting coverage. Age-cohort-specific forecasting was based on incidents with documented patient age. Records with missing patient age were not assigned to age cohorts; instead, projected known-age incidents were scaled to approximate total EMS system volume using the empirically observed relationship between known-age incidents and total reported incidents.

For each historical period, we calculated the known-age share as the proportion of total reported incidents with documented patient age. Projected known-age incident counts were aggregated into annual totals and then adjusted using the observed known-age share to estimate total annual EMS incident volume. This approach preserved interpretable age-specific utilization patterns while accounting for the fact that some EMS records lacked age documentation.

To address variation in reporting coverage over time, the primary adjusted model incorporated reporting-quality variables as exogenous inputs. These variables included the aggregate reporting percentage and the reporting coverage ratio, which were used to account for changes in the completeness and composition of statewide EMS reporting. Projected known-age events were scaled to total system volume using the average known-age share relative to externally reported total incidents, including both digital and aggregate counts.

COVID-era disruption was explicitly modeled using a binary indicator variable in the SARIMAX exogenous inputs. The indicator was coded at the calendar-year level (2020–2021 = 1; all other years = 0), allowing the model to account for temporary deviations in EMS incident patterns during the COVID-19 period without excluding those observations from analysis. This approach preserved the full time series while reducing the influence of pandemic-related disruptions on long-term forecasts.

### Stage 2: Workforce FTE Estimation

In Stage 2, projected EMS incident volumes were converted into estimated EMT and paramedic workforce requirements using a workload-based FTE framework. EMSTARS staffing variables were used to estimate average EMT and paramedic staffing per incident, and incident duration (hours) was used to quantify personnel time per response. Projected annual incident counts were combined with these inputs to estimate total annual workload hours for EMTs and paramedics.

The following operational assumptions were applied:

- Unit Hour Utilization (UHU): 0.35
- Staffing multiplier: 3.5 FTEs per 24-hour position
- Hours per FTE: 8,760 ÷ 3.5 ≈ 2,503 productive hours/year
- Average EMT staffing per incident: 0.6471
- Average paramedic staffing per incident: 1.2142
- Average incident duration: 0.8610 hours

Staffing ratios and duration values were derived from 2017–2025 baseline EMS data. Projected annual incidents were obtained from the forecasting pipeline and used to estimate future staffing requirements.

Let I denote projected annual incidents, E and P the average EMT and paramedic staffing per incident, D the average incident duration (hours), UHU the target unit hour utilization, and H annual productive hours per FTE. Annual workload hours were calculated as:

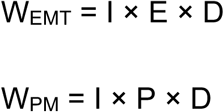

where W_EMT_ and W_PM_ represent total annual workload hours for EMTs and paramedics.

To account for non-response time, workload hours were adjusted using UHU, defined as the proportion of unit time spent on incident response. Required capacity hours were calculated as

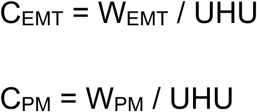

where C_EMT_ and C_PM_ represent utilization-adjusted annual capacity hours for EMTs and paramedics. While a target UHU of 0.35 was specified in the workforce planning assumption to reflect a sustainable operating level, there is no known industry-accepted metric for this measure. As reported by an industry benchmarking initiative, self-described high-performing systems agencies report a UHU between 0.228 and 0.476 (13). This value accounts for non-incident activities, demand variability, recovery time, and system readiness, and was applied as a planning parameter given the absence of a standardized statewide benchmark.

Utilization-adjusted capacity hours were converted to FTE requirements by dividing by annual productive hours per FTE:

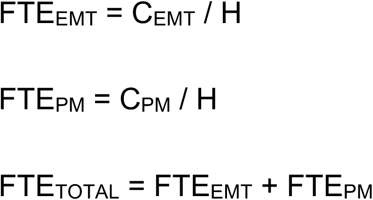

where H = 2,503 productive hours per FTE. Workforce projections were calculated through 2035 and expressed relative to baseline, assuming stable staffing patterns, incident duration, utilization, and productivity.

### Sensitivity Analysis

Sensitivity analyses were conducted to assess the robustness of projected EMS incident volume and associated workforce requirements to alternative modeling assumptions. These analyses evaluated sensitivity to the historical analytic window, reporting-completeness adjustment, missing-age handling, and COVID-era disruption. The primary adjusted model used 2017–2025 data and incorporated reporting-completeness covariates (aggregate reporting percentage and reporting coverage ratio), a COVID-disruption indicator, and scaling procedures to account for missing-age incidents and variation in statewide reporting coverage.

As a sensitivity analysis, we re-estimated the adjusted model using a restricted 2019–2025 analytic window, selected because aggregate-only reporting accounted for less than 10% of total statewide incidents, reflecting improved reporting completeness and reduced reliance on adjustment procedures. The sensitivity model retained the same SARIMAX specification, exogenous variables, COVID-disruption adjustment, known-age cohort forecasting framework, and incident-scaling methodology used in the primary adjusted model. Results were compared with the primary model by examining projected annual EMS incident volume, age-cohort-specific demand patterns, EMS utilization trends, and resulting EMT, paramedic, and total FTE projections through 2035. Consistency between models was interpreted as evidence of robustness to assumptions regarding the historical analytic window and reporting completeness.

Analyses were conducted using Python 3.9. SARIMAX modeling was performed using statsmodels, and data management and aggregation were handled using pandas and NumPy.

## Results

### Population Growth and Aging

Florida’s population grew from 20.5 million in 2017 to 23.3 million in 2025 and is projected to reach 25.8 million by 2035 (Figure 1). Population growth is accompanied by an accelerating aging trend. Adults aged 60 years and older comprised 26.0% of the state in 2017; by 2035, they are projected to represent 31.2% and account for 61.6% of all EMS incidents. The 80+ year-old cohort is the fastest-growing segment, increasing by 45.8% between 2025 and 2035. Table 1 summarizes key population, incident, and workforce metrics across all four reporting years.

**Figure 1.**
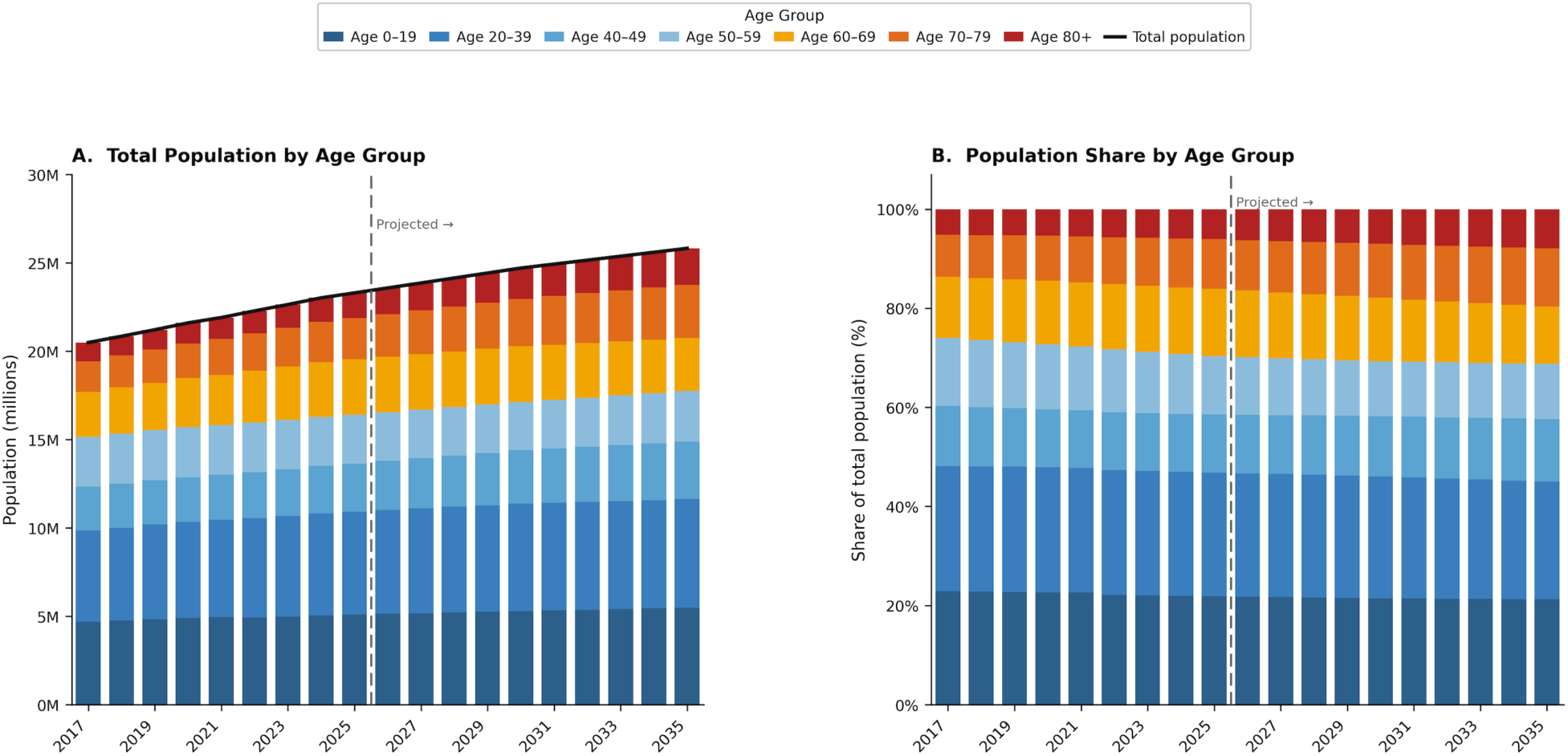
Florida population by age group, 2017–2035. (A) Total population in millions by age group. (B) Percentage distribution of the population by age group. The vertical dashed line indicates the transition from observed to projected values. Figure 1 Footnote: Population estimates (2017-2025) and projections (2026-2035) from the University of Florida Bureau of Economic and Business Research (BEBR). Intermediate-year values were linearly interpolated between available BEBR projection points. Dashed vertical line denotes the transition from observed estimates to projected values Panel A: absolute population by age group (millions) overlaid line indicates total state population. Panel B: Proportional age-group composition.

**Table 1.** Summary of Florida population, EMS incident volume, and estimated workforce requirements, 2017-2035.

Footnote: Projected values for 2030 and 2035 were derived using a SARIMAX forecasting model with BEBR population inputs.
| Metric | 2017 | 2025 | 2030 (proj.) | 2035 (proj.) |
| --- | --- | --- | --- | --- |
| Florida Population |  |  |  |  |
| Total population (millions) | 20.5 | 23.3 | 24.6 | 25.8 |
| Adults ≥60 years (millions) | 5.3 | 6.9 | 7.5 | 8.1 |
| Adults ≥60 as % of population | 26.0% | 29.7% | 30.4% | 31.2% |
| Adults ≥80 years (millions) | 0.9 | 1.4 | 1.8 | 2.1 |
| EMS Incident Volume |  |  |  |  |
| Total annual incidents (millions) | 4.10 | 5.22 | 6.49 | 7.76 |
| Change from 2025 baseline | — | Baseline | +1.27M<br>(+24.4%) | +2.54M<br>(+48.8%) |
| Incidents, adults ≥60 years (millions) | — | 3.08 | 3.92 | 4.78 |
| Adults ≥60 as % of all incidents | — | 59.0% | 60.4% | 61.6% |
| EMS Workforce Requirements |  |  |  |  |
| Total FTE requirement | 7,504 | 9,542 | 11,866 | 14,195 |
| Additional FTEs vs. 2025 baseline | — | Baseline | +2,324<br>(+24.4%) | +4,654<br>(+48.8%) |

**Table 2.**
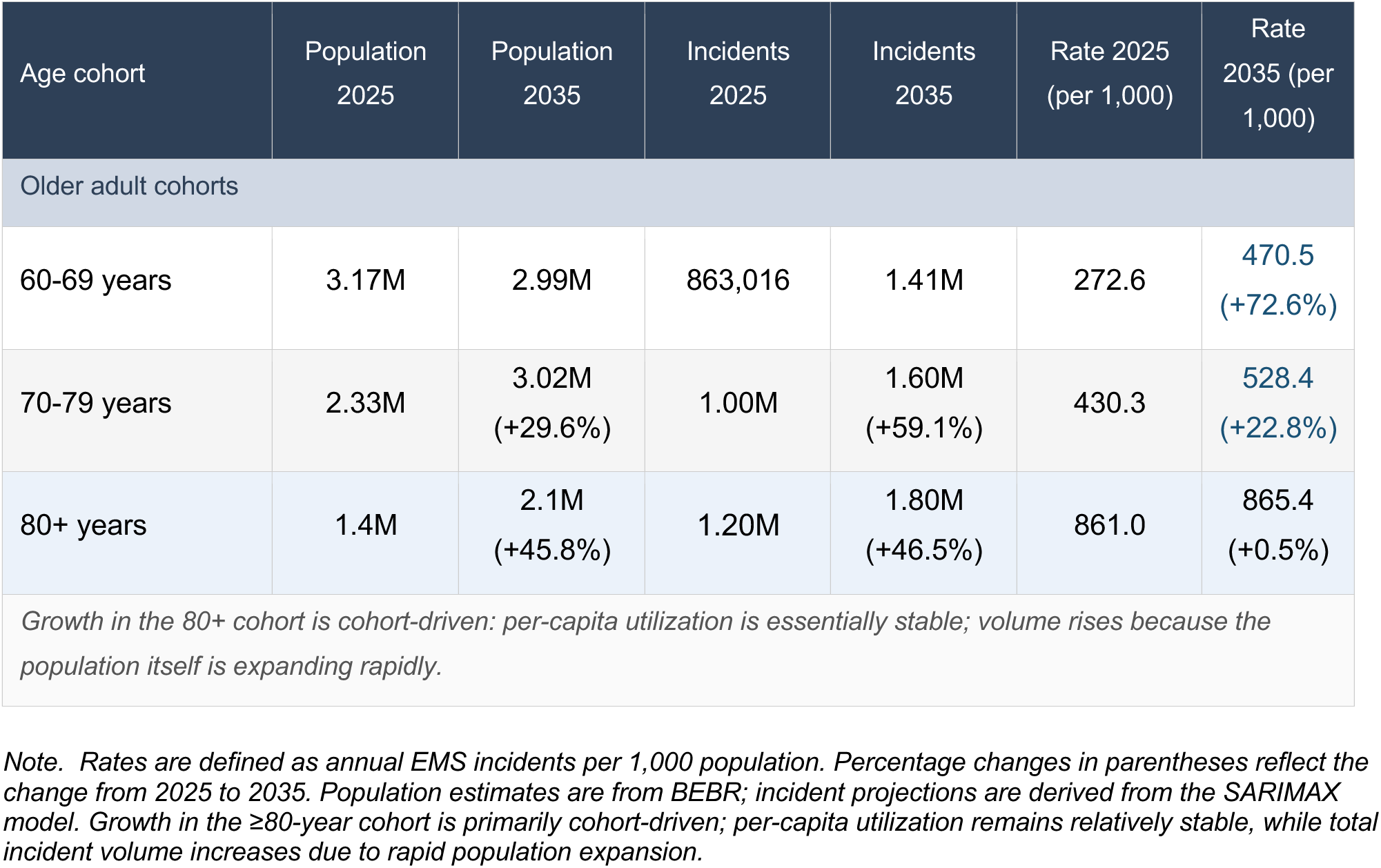
Age cohort population, EMS incident volume, and utilization rates, 2025 and 2035.

| Age cohort | Population 2025 | Population 2035 | Incidents 2025 | Incidents 2035 | Rate 2025 (per 1,000) | Rate 2035 (per 1,000) |
| --- | --- | --- | --- | --- | --- | --- |
| Older adult cohorts |  |  |  |  |  |  |
| 60-69 years | 3.17M | 2.99M | 863,016 | 1.41M | 272.6 | 470.5<br>(+72.6%) |
| 70-79 years | 2.33M | 3.02M<br>(+29.6%) | 1.00M | 1.60M<br>(+59.1%) | 430.3 | 528.4<br>(+22.8%) |
| 80+ years | 1.4M | 2.1M<br>(+45.8%) | 1.20M | 1.80M<br>(+46.5%) | 861.0 | 865.4<br>(+0.5%) |
| Growth in the 80+ cohort is cohort-driven: per-capita utilization is essentially stable; volume rises because the population itself is expanding rapidly. |  |  |  |  |  |  |

*Note. Rates are defined as annual EMS incidents per 1,000 population. Percentage changes in parentheses reflect the change from 2025 to 2035. Population estimates are from BEBR; incident projections are derived from the SARIMAX model. Growth in the ≥80-year cohort is primarily cohort-driven; per-capita utilization remains relatively stable, while total incident volume increases due to rapid population expansion.*

### EMS Incident Volume

Annual EMS incidents are projected to increase by 48.8% from the 2025 baseline, reaching 7.76 million by 2035 (Figure 2). Growth is heavily concentrated among older adults. By 2035, adults aged 60 years and older will represent nearly one-third of Florida’s population, yet account for nearly two-thirds of all EMS incidents.

**Figure 2.**
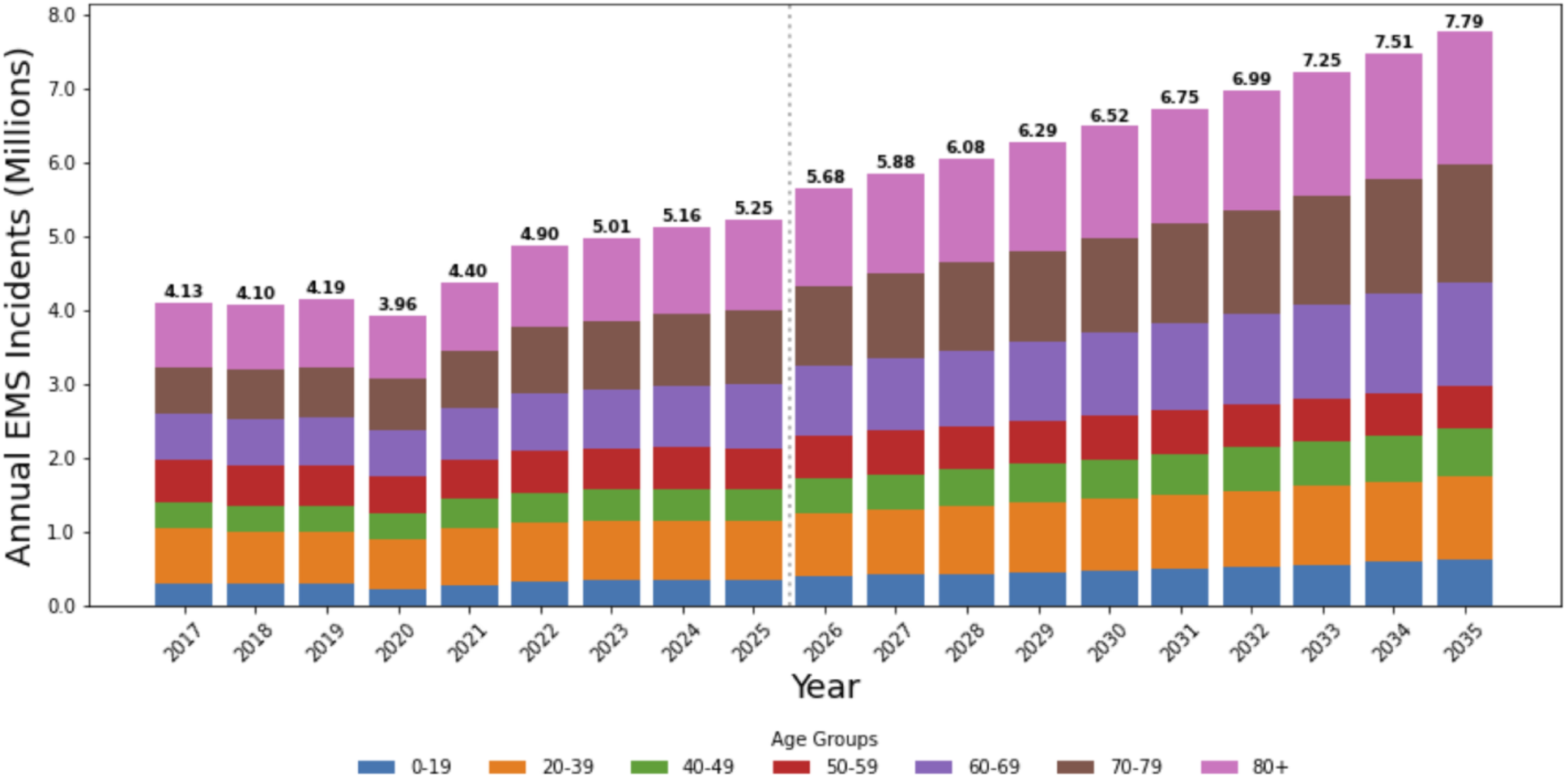
Annual EMS incidents (millions) by age group, 2017–2035. Note. The vertical dashed line marks the transition from historical observed data (through 2025) to projected values (2026–2035).

### Age-Specific Utilization

Incident growth among older adults reflects distinct dynamics across age cohorts. Among adults aged 60–69, projected incident growth is driven primarily by increasing per-capita utilization despite a modest decline in population size. Among adults aged 70–79, both population growth and increasing per-capita utilization contribute to projected incident growth. Among adults aged 80 and older, per-capita utilization is essentially stable; projected incident growth is driven entirely by cohort expansion.

Age-specific utilization increased with age throughout the study period and the projection horizon (Figure 3). The highest rates were consistently observed in the 80+ year cohort. However, the steepest projected increase occurs among adults aged 60-69 years, whose utilization rate is projected to be 135.4% higher than that of the 50-59 year cohort by 2035, compared with 20.0% higher in 2017. These findings indicate that future EMS demand is not explained by population growth alone.

**Figure 3.**
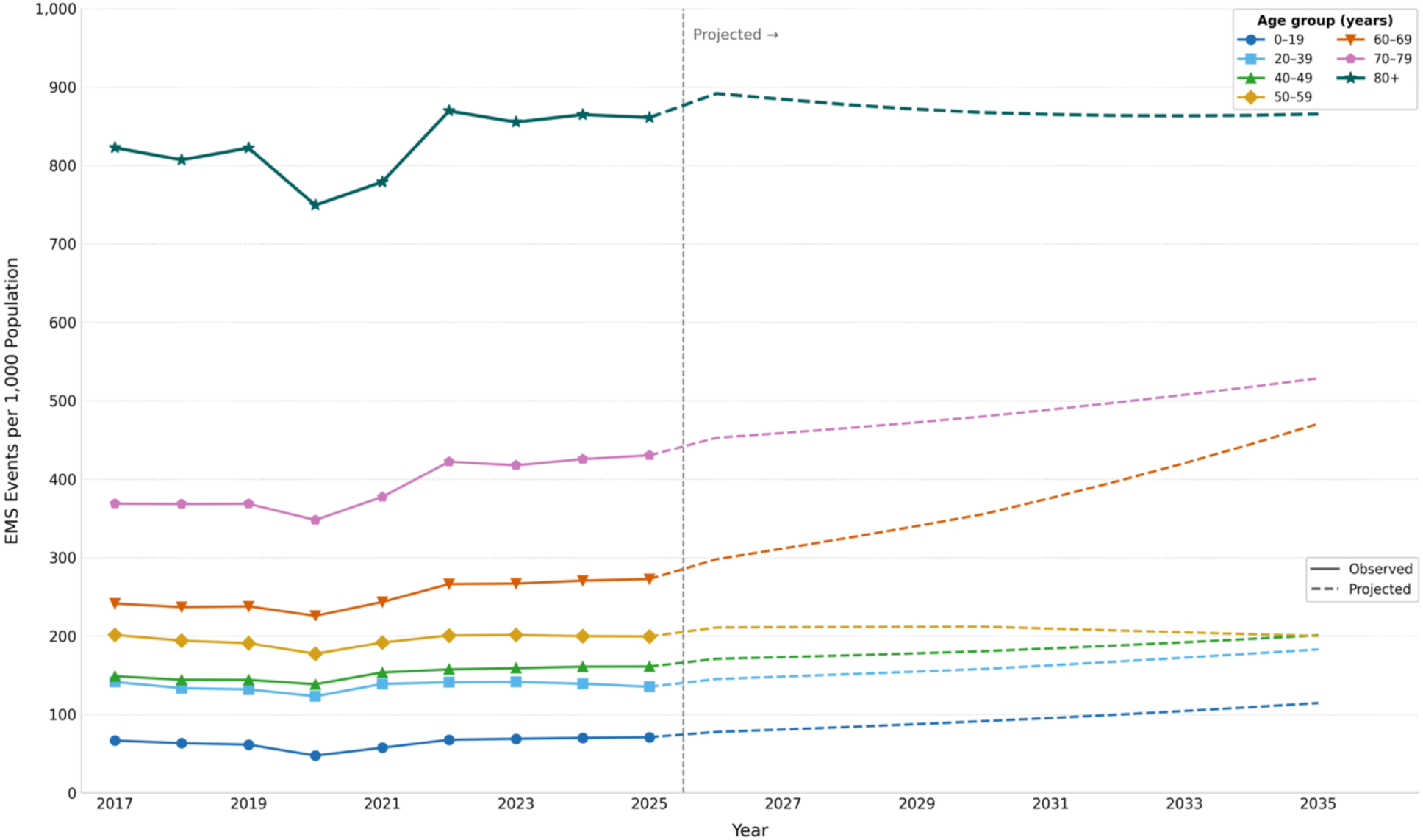
EMS Utilization Rates per 1,000 Population by Age Group, 2017-2035. Note. Utilization rates represent annual EMS incidents divided by population within each age group, expressed per 1,000 population.

### Estimated Workforce Requirements

Estimated FTE requirements follow the projected incident curve (Figure 4). Total requirements are projected to rise from 9,542 FTEs in 2025 to 14,195 FTEs by 2035, an increase of 4,654 FTEs (48.8%) above the 2025 baseline. These projections reflect demand-based net requirements under current staffing assumptions and do not include attrition.

**Figure 4.**
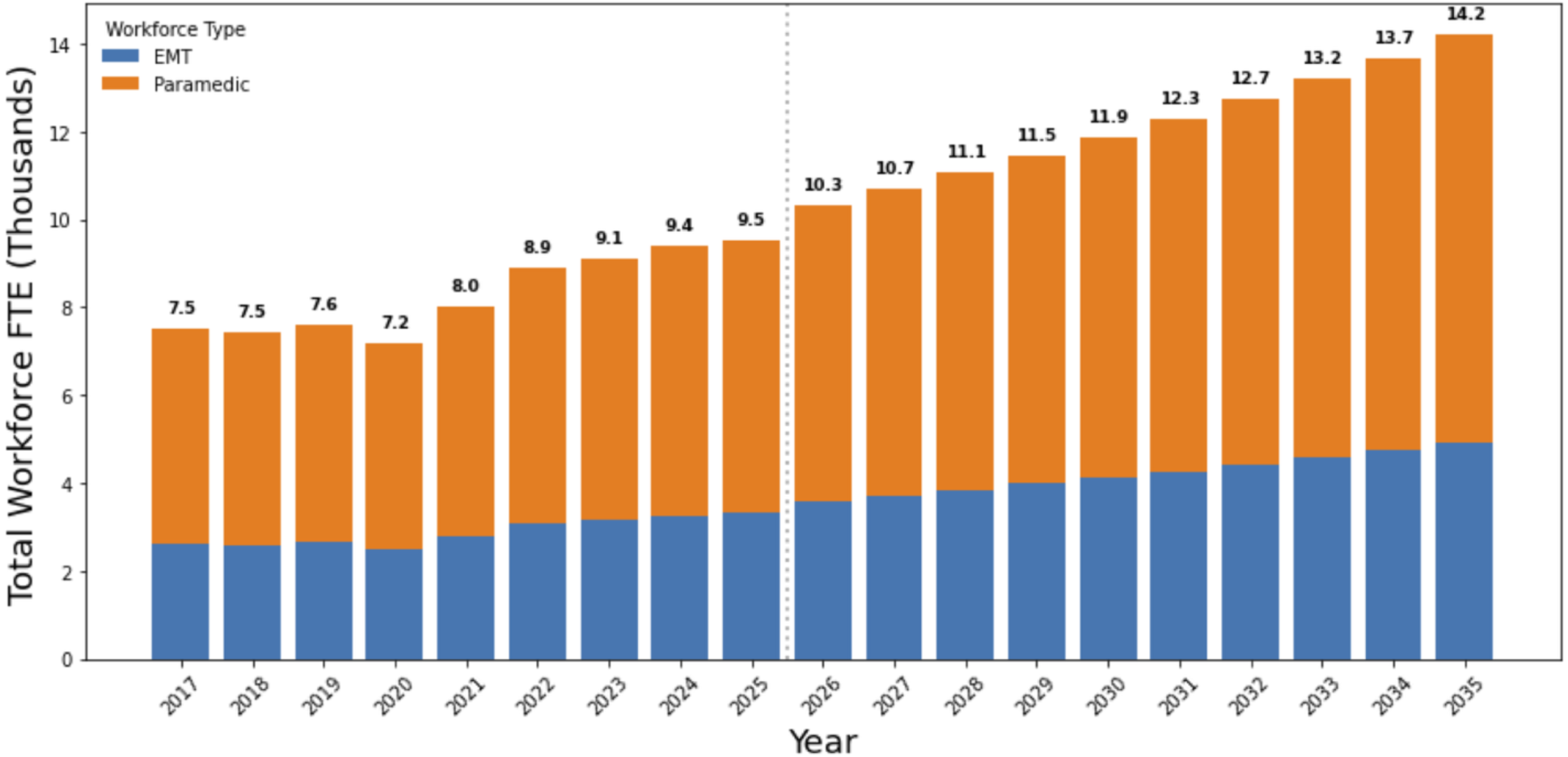
Actual & estimated EMT and paramedic workforce FTE requirements, Florida, 2017-2035. Note. Values represent estimated optimized workforce full-time equivalent (FTE) requirements derived from EMS incident volume, observed EMSTARS staffing configurations, average incident duration, target unit-hour utilization (UHU = 0.35), and 2,503 annual productive hours per FTE. The vertical dashed line indicates the transition from historical/model-fitting years to projected values.

Collectively, these results demonstrate that projected EMS demand in Florida is driven by the interplay of statewide population growth, rapid growth in the oldest age cohorts, and rising age-specific utilization, particularly among adults in their 60s, as the baby boom cohort moves through the young-old years.

## Discussion

Emergency medical services constitute a foundational element of the public health infrastructure, serving as the primary prehospital interface between community health needs and acute care delivery. The ability to project EMS service demand and associated clinician workforce requirements with reasonable precision is, therefore, essential to the evidence-based planning that state and local agency leaders and policymakers require to sustain system capacity as population aging accelerates. Florida’s experience is particularly instructive in this regard. As home to one of the nation’s largest concentrations of adults aged 80 and older and a rapidly expanding oldest-old cohort, Florida is confronting at scale the demographic conditions that other states will face within the coming decade. The forces driving projected EMS demand growth here, accelerating cohort expansion, sustained retiree in-migration, and a growing imbalance between older residents and the working-age population, represent a generalizable case study whose implications extend well beyond a single state (14, 15).

This study makes two distinct methodological contributions. First, to our knowledge, it is the first to apply SARIMAX-based demand forecasting to statewide EMS data in conjunction with validated age-cohort population projections to generate state-level workforce requirements. Second, the model was trained on nine years of historical data from January 1, 2017 through December 31, 2025, encompassing a pre-pandemic baseline, the COVID-19 disruption and recovery, and the post-pandemic utilization trajectory, thus anchoring projections to the most temporally complete dataset currently available.

The most consequential finding for both clinical practice and workforce planning is the extent to which projected demand is concentrated in the oldest age cohorts. Adults aged 80 and older are projected to account for a disproportionate share of total EMS incidents through 2035, despite comprising a relatively small share of Florida’s population. Analysis of age-specific utilization rates shows that per capita contact rates in this cohort have remained essentially stable over time. The projected surge in absolute incident volume is therefore driven not by rising individual-level utilization, but by cohort expansion as a growing share of Florida’s population ages into the oldest-old age range.

This distinction carries an important policy implication. Demand growth driven by population aging rather than rising rates of illness per person can be challenging to reduce through individual-level interventions or utilization management programs that target behavioral change. This is not to say, however, that such interventions should not be explored, particularly considering that the demand surge extends well beyond the oldest old. Adults aged 60–69 are projected to experience increasing EMS utilization despite a modest decline in cohort size, whereas adults aged 70–79 experience growth in both cohort size and per-capita utilization as the leading edge of the baby boom moves through the young-old years — a compounding dynamic that distinguishes this period from prior demographic transitions. These trends are consistent with published evidence that EMS utilization among the young old reflects the accumulating burden of multiple chronic conditions and functional decline characteristic of those in their 60s and 70s (5, 16–18). For EMS systems, the implication is that demand growth is not a single peak associated with the oldest cohort, but a sustained increase across the full range of older adult ages.

These projections build on well-established literature documenting older adults’ disproportionate use of EMS and the clinical complexity of their presentations. National studies consistently show that older age is associated with substantially higher transport rates, a greater likelihood of emergency department evaluation, and increased hospitalization after EMS contact (9, 10, 17). The drivers identified in that literature which include falls, chronic disease exacerbations, dementia-related episodes, and unmet social and functional support needs, are also the conditions whose prevalence rises as populations move into the oldest age categories (19–21). What the present study adds is a prospective, statewide quantification of the implications of these well-documented clinical patterns for system capacity and workforce requirements as demographic change scales them to a new magnitude.

The workforce projections reported here reflect demand-based net requirements and exclude attrition from retirement, voluntary turnover, or career transitions. National data indicate rising burnout rates, with higher prevalence among paramedics than among EMTs (22). Florida-specific data documents persistent staffing gaps, elevated turnover intentions, and operational pressures that both reflect and accelerate attrition (23, 24). When attrition is incorporated alongside projected demand growth, gross recruitment will exceed the net FTEs estimate shown above. Comprehensive workforce planning requires pairing of demand-side projections with supply-side modeling that incorporates pipeline capacity, graduation rates, and retention under realistic scenarios.

The projected workforce growth of approximately 48.8% by 2035 has fiscal and operational implications that warrant immediate attention from agency leaders and policymakers. States considering community paramedicine programs or investing in EMS education would benefit from similarly rigorous future demand modeling. Florida’s experience will provide a directly applicable template for the many states that will face comparable conditions within the next decade.

## Limitations

This study has several limitations. Incident forecasts assume that historical age-specific EMS utilization patterns provide a reasonable basis for projecting demand through 2035. Adjustments were made for reporting completeness, missing age data, and COVID-era disruption. Although the model accounts for population growth and aging, it does not explicitly incorporate future changes in population health, health care access, medical technology, EMS scope of practice, payer policy, or care delivery models that could alter utilization.

Workforce estimates assume that historical staffing configurations, average incident duration, and operational efficiency persist over the projection horizon. These inputs were derived from EMSTARS data and applied to projected incident volumes; however, future changes in deployment models, response protocols, transport practices, documentation requirements, or system efficiency were not modeled and may affect staffing needs.

The COVID-19 adjustment captures observed disruption during 2020–2021 but does not account for future pandemics, disasters, policy interventions, or system changes (e.g., expanded community paramedicine) that could shift demand. Despite adjustments for missing age and reporting variation in the modeling, some residual misclassification or reporting bias may remain.

Finally, the analysis reflects statewide patterns in Florida and does not assess substate variation. Workforce estimates should be interpreted as demand-based planning targets under specified assumptions. Future studies should examine trends in incident and workforce estimates at the substate level to inform local agency-specific workforce planning.

## Conclusions

The growth and aging of Florida’s population will drive a substantial and sustained increase in EMS demand over the next decade. Adults aged 60 and older, particularly those 80 and older, will account for a disproportionate and growing share of EMS incidents under any plausible demographic scenario. These forces will place considerable pressure on the EMS workforce capacity under current staffing and operational assumptions, and the window for proactive workforce planning is narrowing.

Florida’s experience is at the leading edge of a demographic transformation that will ultimately reach every state in the nation. With one of the nation’s largest concentration of adults aged 80 and older, accelerating retiree in-migration, and a rapidly growing oldest-old cohort, Florida is confronting, at scale, what other states will face in the coming decade. The demographic forces documented here are national in scope; their arrival in other states is a matter of timing, not uncertainty.

Addressing the workforce demands projected in this study will require sustained, multi-level responses. Expanding models such as community paramedicine, mobile integrated health, telehealth-assisted triage, and age-friendly emergency care may be meaningful mitigation strategies; however, the evidence base for their impact at scale remains limited and warrants further investigation. Structural interventions to strengthen EMS education pipeline capacity, recruitment, and retention will also be necessary complements to any demand-side mitigation. The findings presented here suggest that these responses must be planned and resourced proactively, before demand growth outpaces the systems designed to meet it.

## Data Availability

All data produced in the present study are available upon reasonable request to the authors

## ACKNOWLEDGEMENTS

The authors thank Tanner Pasman and Ruth Esther Challa, MD, MPH, of the University of South Florida College of Public Health, for their contributions to the early stages of this research, including their assistance with the literature search.

## Disclosures

### DECLARATION OF GENERATIVE AI IN SCIENTIFIC WRITING

During the preparation of this work, BJM and LJP used Claude Sonnet 4.6 (Anthropic, San Francisco, CA) to assist with reference identification, citation formatting, and manuscript editing. After using this tool, the authors reviewed and edited the content as needed. The authors take full responsibility for the content of this publication.

### DATA SHARING STATEMENT

The EMS incident data used in this study were obtained from the Florida Emergency Medical Services Tracking and Reporting System (EMSTARS) under a data use agreement with the Florida Department of Health and cannot be made publicly available by the authors. Population projection data are publicly available from the University of Florida Bureau of Economic and Business Research (BEBR) at https://bebr.ufl.edu. Analytic code and supplemental methods documentation are available from the corresponding author upon reasonable request.

### AUTHORSHIP STATEMENT

All authors participated in substantial contributions to this project. BJM, ML, HM, and LJP conceived and/or helped design the study; BJM, ML, and MAO acquired and analyzed the data. All authors drafted the manuscript and/or contributed substantially to its revision for important intellectual content. All authors provided final approval of the version to be published and agreed to be accountable for all aspects of the work.

## References

1. Vespa J, Medina L, Armstrong DM. Demographic Turning Points for the United States: Population Projections for 2020 to 2060. Current Population Reports, P25-1144. Washington, DC: U.S. Census Bureau; 2020. Available from: https://www.census.gov/library/publications/2020/demo/p25-1144.html

2. Ortman JM, Velkoff VA, Hogan H. An Aging Nation: The Older Population in the United States. Current Population Reports, P25-1140. Washington, DC: U.S. Census Bureau; 2014. Available from: https://www.census.gov/content/dam/Census/library/publications/2014/demo/p25-1140.pdf

3. U.S. Department of Health and Human Services. 2023 Profile of Older Americans. Washington, DC: Administration for Community Living; 2024. Available from: https://acl.gov/sites/default/files/Profile%20of%20OA/ACL_ProfileOlderAmericans2023_508.pdf

4. Khan HTA, Addo KM, Findlay H. Public health challenges and responses to the growing ageing populations. Public Health Challenges. 2024;3(3):e213. doi:10.1002/puh2.213

5. Watson KB, Wiltz JL, Nhim K, Kaufmann RB, Thomas CW, Greenlund KJ. Trends in multiple chronic conditions among US adults, by life stage, Behavioral Risk Factor Surveillance System, 2013–2023. Prev Chronic Dis. 2025;22:240539. doi:10.5888/pcd22.240539

6. Ritchie CS, Leff B. Home-based care reimagined: a full-fledged health care delivery ecosystem without walls. Health Aff. 2022;41(5):689–95. doi:10.1377/hlthaff.2021.01011

7. Lochner KA, Goodman RA, Posner S, Parekh A. Multiple chronic conditions among Medicare beneficiaries: state-level variations in prevalence, utilization, and cost, 2011. Medicare Medicaid Res Rev. 2013;3(3):E1–19. doi:10.5600/mmrr.003.03.b02

8. Platts-Mills TF, Leacock B, Cabañas JG, Shofer FS, McLean SA. Emergency medical services use by the elderly: analysis of a statewide database. Prehosp Emerg Care. 2010;14(3):329–33. doi:10.3109/10903127.2010.481759

9. Shah MN, Bazarian JJ, Lerner EB, Fairbanks RJ, Barker WH, Auinger P, et al. The epidemiology of emergency medical services use by older adults: an analysis of the National Hospital Ambulatory Medical Care Survey. Acad Emerg Med. 2007;14(5):441–7. Available from: https://pubmed.ncbi.nlm.nih.gov/17456555/

10. Jones CMC, Wasserman EB, Li T, Amidon A, Abbott M, Shah MN. The effect of older age on EMS use for transportation to an emergency department. Prehosp Disaster Med. 2017;32(3):261–8. doi:10.1017/S1049023X17000036

11. Zaphir JS, Murphy KA, MacQuarrie AJ, Stainer MJ. Understanding the role of cognitive load in paramedical contexts: a systematic review. Prehosp Emerg Care. 2025;29(2):101–14. doi:10.1080/10903127.2024.2370491

12. Evans CS, Platts-Mills TF, Fernandez AR, Grover JM, Cabanas JG, Patel MD, et al. Repeated emergency medical services use by older adults: analysis of a comprehensive statewide database. Ann Emerg Med. 2017;70(4):506–15.e3. doi:10.1016/j.annemergmed.2017.03.058

13. Simpson K, Pierson J. Where should you start when it comes to benchmarking? The AIMHI benchmarking project focuses on high-performance, high value operational measures. EMS1. December 30, 2020. https://www.ems1.com/administration/articles/where-should-you-start-when-it-comes-to-benchmarking-V0MsK9HuzYPoTzRl/

14. Maestas N, Mullen KJ, Powell D. The effect of population aging on economic growth, the labor force, and productivity. Am Econ J Macroecon. 2023;15(2):306–32. doi:10.1257/mac.20190196

15. Congressional Budget Office. The Demographic Outlook: 2025 to 2055. Washington, DC: Congressional Budget Office; 2025. Report No.: 61164. Available from: https://www.cbo.gov/publication/61164

16. Günay Tuzcu G, Ekşi A. Determinants of emergency medical services utilization among older adults: a comprehensive scoping review. Geriatr Nurs. 2025;66(Pt B):103623. doi:10.1016/j.gerinurse.2025.103623

17. Duong HV, Herrera LN, Moore JX, Donnelly J, Jacobson KE, Carlson JN, et al. National characteristics of emergency medical services responses for older adults in the United States. Prehosp Emerg Care. 2018;22(1):7–14. doi:10.1080/10903127.2017.1347223

18. Burnett SJ, Alianell T, Bitnok O, Ebersole K, Nuruddin B, Butler S, et al. Social determinants of health and emergency medical services: a scoping review. Prehosp Emerg Care. 2026;30(2):181–94. doi:10.1080/10903127.2025.2468796

19. Faul M, Stevens JA, Sasser SM, Alee L, Deokar AJ, Kuhls DA, et al. Older adult falls seen by emergency medical service providers: a prevention opportunity. Am J Prev Med. 2016;50(6):719–26. doi:10.1016/j.amepre.2015.12.011

20. Freedman VA, Spillman BC. Disability and care needs among older Americans. Milbank Q. 2014;92(3):509–41. doi:10.1111/1468-0009.12076

21. Martínez B, Aranda MP, Sanko S, Aguilar I, Vega WA. Older adult frequent 9-1-1 callers for emergency medical services in a large metropolitan city: individual- and system-level considerations. J Emerg Med. 2023;65(6):e522–e30. doi:10.1016/j.jemermed.2023.07.006

22. Powell JR, Gage CB, Crowe RP, Rush LJ, MacEwan SR, Dixon G, et al. National evaluation of emergency medical services clinician burnout and workforce-reducing factors. JACEP Open. 2025;6(1):100024. doi:10.1016/j.acepjo.2024.100024

23. Moeller BJ, Lozano M. Crisis on the Frontlines: Recruitment and Retention Challenges in EMS. Tampa, FL: University of South Florida, Morsani College of Medicine, Florida Center for EMS; August 2025. doi:10.5038/XDPT2458

24. Lozano M, Moeller BJ, Tobin A. Vital Signs: Findings from Florida’s 2024 EMS Workforce Statewide Survey. Tampa, FL: University of South Florida, Morsani College of Medicine, Florida Center for EMS; 2025. doi:10.5038/UCID5504

